# Evaluation and clinical implications of the time to a positive results of antigen testing for SARS-CoV-2

**DOI:** 10.1101/2021.06.09.21258157

**Authors:** Yusaku Akashi, Yoshihiko Kiyasu, Yuto Takeuchi, Daisuke Kato, Miwa Kuwahara, Shino Muramatsu, Atsuo Ueda, Shigeyuki Notake, Koji Nakamura, Hiroichi Ishikawa, Hiromichi Suzuki

## Abstract

Antigen tests for severe acute respiratory coronavirus 2 sometimes show positive lines earlier than their specified read time, although the implication of getting the results at earlier time is not well understood. This study aimed to evaluate the clinical utility of an antigen test by evaluating the time period to get positive results and by comparing the test sensitivity with that of a digital immunoassay (DIA) test.

We prospectively collected additional nasopharyngeal samples from patients who had already tested positive for SARS-CoV-2 by reverse transcription PCR. The additional swab was used for an antigen test, QuickNavi™-COVID19 Ag, and the time periods to get positive results were measured. The sensitivity of QuickNavi™-COVID19 Ag was also compared with that of a DIA.

In 84 of 96 (87.5%) analyzed cases, the results of QuickNavi™-COVID19 Ag were positive. The time to obtain positive results was 15.0 seconds in median (inter quartile range: 12.0-33.3, range 11-736), and was extended in samples with higher cycle thresholds (Ct) (*p*<0.001). Positive lines appeared within a minute in 85.7% of cases and within 5 minutes in 96.4%. The sensitivities of QuickNavi™-COVID19 Ag and the DIA were 87.5% (95% confident interval [CI]: 79.2%-93.4%) and 88.6% (95%CI: 75.4%- 96.2%), respectively. Their results were concordant in 90.9% of cases, with discrepancies present only in cases with Ct values >32.

QuickNavi™-COVID19 Ag immediately showed positive results in most cases, and the time to a positive reaction may have indicated the viral load. In addition, the sensitivity of the test was comparable to the DIA.

## Introduction

The global pandemic caused by severe acute respiratory coronavirus 2 (SARS-CoV-2) remains a concern and is unlikely to end soon [1]. Until the pandemic is under control, early diagnosis and screening with effective test modalities are essential strategies to combat the disease [2].

Antigen tests originally had low sensitivity compared to nucleic examinations [3]. Their diagnostic performance has improved over time [4], and previous research has found them beneficial for screening purposes [5] or in low-resource settings [6]. Advantages of antigen tests include the ability to analyze multiple samples simultaneously, the lack of a need for special equipment, and the fast turnaround time (TAT).

Manufacturers of antigen tests recommend that the results be read 10-30 minutes after sample placement [3]. Positive lines sometimes appear before the manufacturer’s specified time, although the clinical implication of an early detection is not well understood. Knowing the time to a positive reaction in clinical samples may help identify a more appropriate read time, which could reduce their overall TAT.

We previously evaluated the diagnostic performance of QuickNavi™-COVID19 Ag (Denka Co., Ltd., Tokyo, Japan), revealing a sensitivity of 86.7% and a specificity of 100% [7], with a read time of 15 minutes. The sensitivity seemed sufficient, although the limit of detection was inferior to that of a rapid immunochromatography, which utilizes a digital immunoassay (DIA) platform [8]. The head-to-head comparison using clinical samples has been awaited to validate their test performances.

In this study, we aimed to clarify the clinical utility of QuickNavi™-COVID19 Ag by evaluating the time periods to get positive results and by comparing its sensitivity with an immunochromatography test, which comes with a DIA reader.

## Methods

Between January 25, 2021, and March 27, 2021, we prospectively evaluated the time necessary for positive reactions in two antigen tests; one is QuickNavi™-COVID19 Ag, which is already on the market with 15 minutes read time, and the other is its improved product (DK20-CoV-8M). Nasopharyngeal samples were obtained from SARS-CoV-2–positive patients after they provided informed consent. The ethics committee of Tsukuba Medical Center Hospital (TMCH) approved the present study (approval number: 2020-033).

### Study sites and patient backgrounds

This study was performed at a drive-through-type PCR center located at TMCH in Tsukuba, Japan. The PCR center performed in-house RT-PCR examinations [7] of residents of the Tsukuba district who were referred from a local public health center and primary care facilities (Figure 1). PCR examination was performed immediately after each nasopharyngeal sampling, and the results were reported to both patients and doctors within a few hours. Physicians then assessed the risk factors and disease severity of PCR-positive patients (Figure 1).

**Figure 1.**
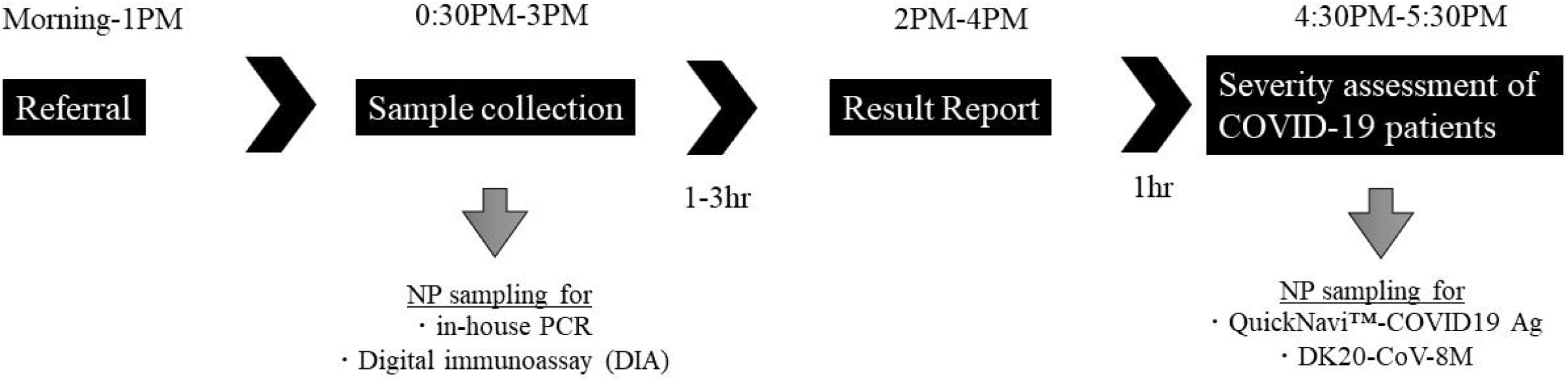
Workflow of a drive-through testing site at Tsukuba Medical Center Hospital. Abbreviation: NP, nasopharynx

### Sample collection and antigen test procedures

In the current study, an additional nasopharyngeal sample was obtained from in-house PCR-positive patients using FLOQSwab™ (Copan Italia S.p.A., Brescia, Italy), using a previously described technique [9]. The swab sample was diluted with specimen buffer provided in a tube, which is included in the test kit, and commonly used for the two antigen tests. The tests were simultaneously performed according to the manufacturers’ instructions (Figure 2), and examiners visually interpreted the results. The required time for positive results was measured with a stopwatch after specimens were added to the test cassette. If the line indicating positivity was not visualized, the results were recorded as negative.

**Figure 2.**
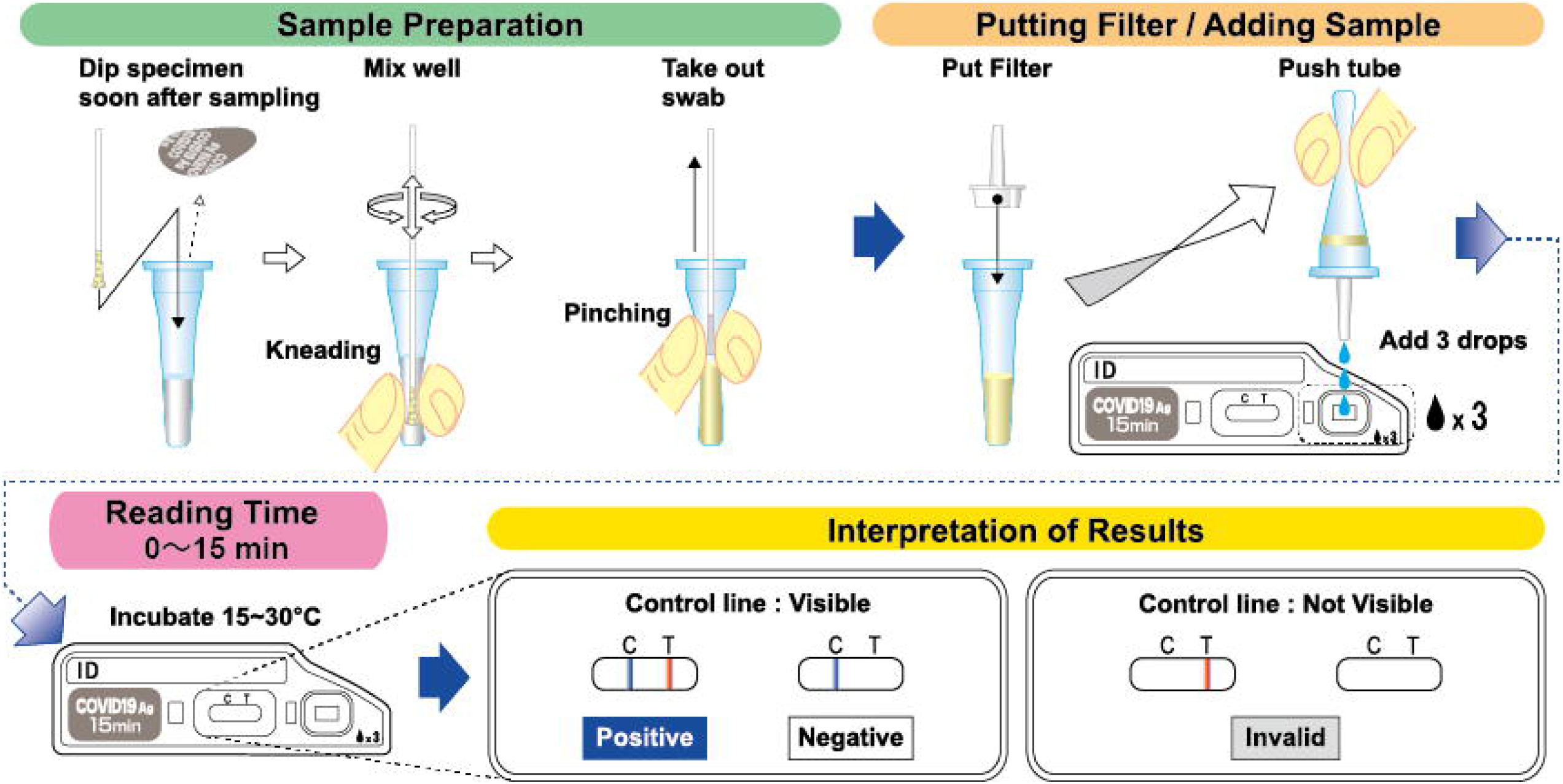
Procedures for DK20-CoV-8M testing.

### Copy numbers and Ct values of PCR examination in comparison with the required time for positive results of antigen test

All the antigen test evaluations were performed on the same day of sampling specimens for RT-PCR examinations. The RT-PCR examinations used nasopharyngeal samples suspended in 3 mL of UTM™ (Copan Italia S.p.A.). After extraction with a magLEAD 6gC (Precision System Science Co., Ltd., Chiba, Japan), the eluted samples were first tested with in-house PCR. Rest of the eluted RNA were stored at − 80 °C and were transferred to Denka Co., Ltd., every week for real-time RT-PCR of SARS-CoV-2. The examination employed the QuantStudio^®^3 Real-Time PCR System (Thermo Fisher Scientific Inc., MA, USA) and a method developed by the National Institute of Infectious Diseases (NIID), Japan [10]. The NIID N/N2 SARS-CoV-2 Standard (Nihon Gene Research Laboratories, Inc., Japan) was used as a control to make a standard curve for the analysis of copy numbers.

### Comparison between QuickNavi™-COVID19 Ag and DIA

During the study period, antigen testing was performed with both QuickNavi™-COVID19 Ag and QuickChaser Auto SARS-CoV-2 (Mizuho Medy Co., Ltd.) at the drive-through-type PCR center. We compared the analytical sensitivity for the detection of SARS-CoV-2 between the two antigen tests. The results of real-time RT-PCR of SARS-CoV-2 were used as the reference standard.

### Statistical analyses

The sensitivity of antigen testing was calculated using the Clopper and Pearson method, with 95% confident intervals (95% CI). Linear regression analysis was used to describe the relationship between two continuous variables; *p*-values <0.05 were considered to represent statistically significant differences. All calculations were conducted using R 4.0.4 (The R Foundation, Vienna, Austria).

## Results

The study included 96 patients who were positive for SARS-CoV-2 by RT-PCR, of whom 52 (54.2%) were symptomatic. The RT-PCR examination detected the N2 gene in all patients and failed to identify the N gene in 11. The median number of viral copies per test and the median Ct values were 360,000 (inter quartile range [IQR]: 22,000-2,300,000) and 28.0 (IQR: 25.0-32.0) for the N gene, and 1,250,000 (IQR: 82,750-20,500,000) and 24.0 (IQR: 19.8-28.0) for the N2 gene. The numbers of cases with an N2 gene Ct of <20, 20-30, and ≥30 were 24, 52, and 20, respectively.

QuickNavi™-COVID19 Ag and DK20-CoV-8M were positive in the same 84 patients; thus, their sensitivities were 87.5% (95%CI: 79.2%-93.4%) when using N2 gene RT-PCR as a reference (Table 1). False negatives occurred in 12 patients, and in all of them the N2 gene Ct values were >32.

**Table 1.** Sensitivities and times to a positive reaction for three antigen tests

|  | Sensitivity<br>(95%CI), % | Median time to a positive<br>reaction [IQR], sec |
| --- | --- | --- |
| QuickNavi™-COVID19 Ag | 87.5 (79.2-93.4) | 15.0 [12.0-33.3] |
| DK20-CoV-8M | 87.5 (79.2-93.4) | 21.0 [17.0-44.3] |
| DIA | 88.6 (75.4-96.2) | - |

CI, confidence interval; IQR, interquartile range; DIA, digital immunoassay
The time to a positive reaction was not available for DIA due to the nature of digital immunoassays. The running time of its reader is 15 minutes.
The RT-PCR of the N2 gene was used as a reference.

### Time required to get positive results for QuickNavi™-COVID19 Ag and DK20-CoV-8M

The median time to get positive results was 15.0 seconds (IQR: 12.0-33.3, range 11-736) with QuickNavi™-COVID19 Ag and 21.0 seconds (IQR: 17.0-44.3, range: 11-736) with DK20-CoV-8M (Table 1).

Positive lines were observed within a minute in 85.7% of samples analyzed with QuickNavi™-COVID19 Ag and in 78.6% of those analyzed with DK20-CoV-8M (Figure 3). In 96.4% of cases, both tests can be interpreted as positive within 5 minutes. The time required to get positive results for the two antigen tests was significantly correlated, as shown in the Supplementary Figure (*p<*0.001). In three cases, more than 5 minutes were required to obtain positive results in both antigen tests; their Ct values were 28, 34, and 38, respectively.

**Figure 3.**
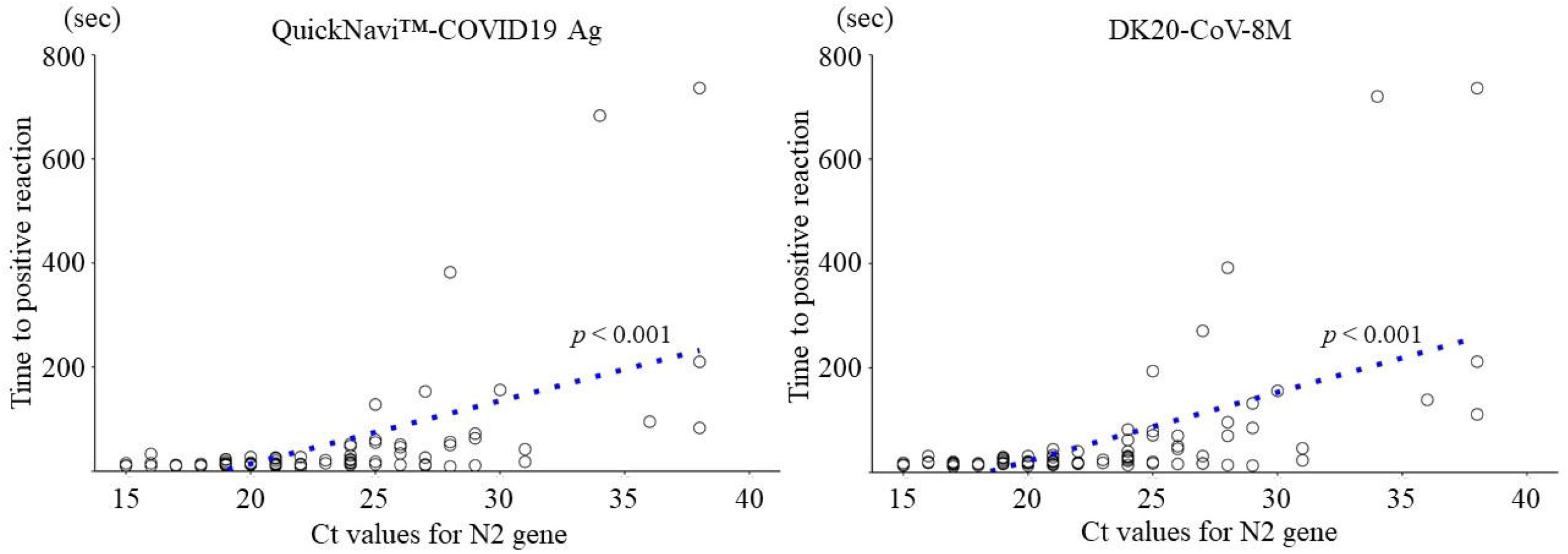
Relationship between the time to get positive results and sample Ct values. Circles indicate cases with positive antigen tests. The results of linear regression analyses are shown as dotted lines, along with their p values.

As depicted in Figure 3, there was a linear association between the time required to get positive results and Ct values (N2 gene) in samples (*p*<0.001).

### Concordance of QuickNavi™-COVID19 Ag with DIA

Of the 96 included patients, 44 provided samples for antigen testing by the DIA, and the results were concordant with those of QuickNavi™-COVID19 Ag in 90.9% of cases (40/44). Both tests exhibited positive results in all samples with N2 gene Ct values ≤31 (Figure 4). Discordant results were observed in four samples: in two, with Ct values of 36 and 38, respectively, QuickNavi™-COVID19 Ag was positive while the DIA was negative, while in the other two, with Ct values of 33 and 34, respectively, QuickNavi™-COVID19 Ag was negative and the DIA was positive. Neither antigen test could detect the virus in three samples with Ct values of 34 and 36, respectively.

**Figure 4.**
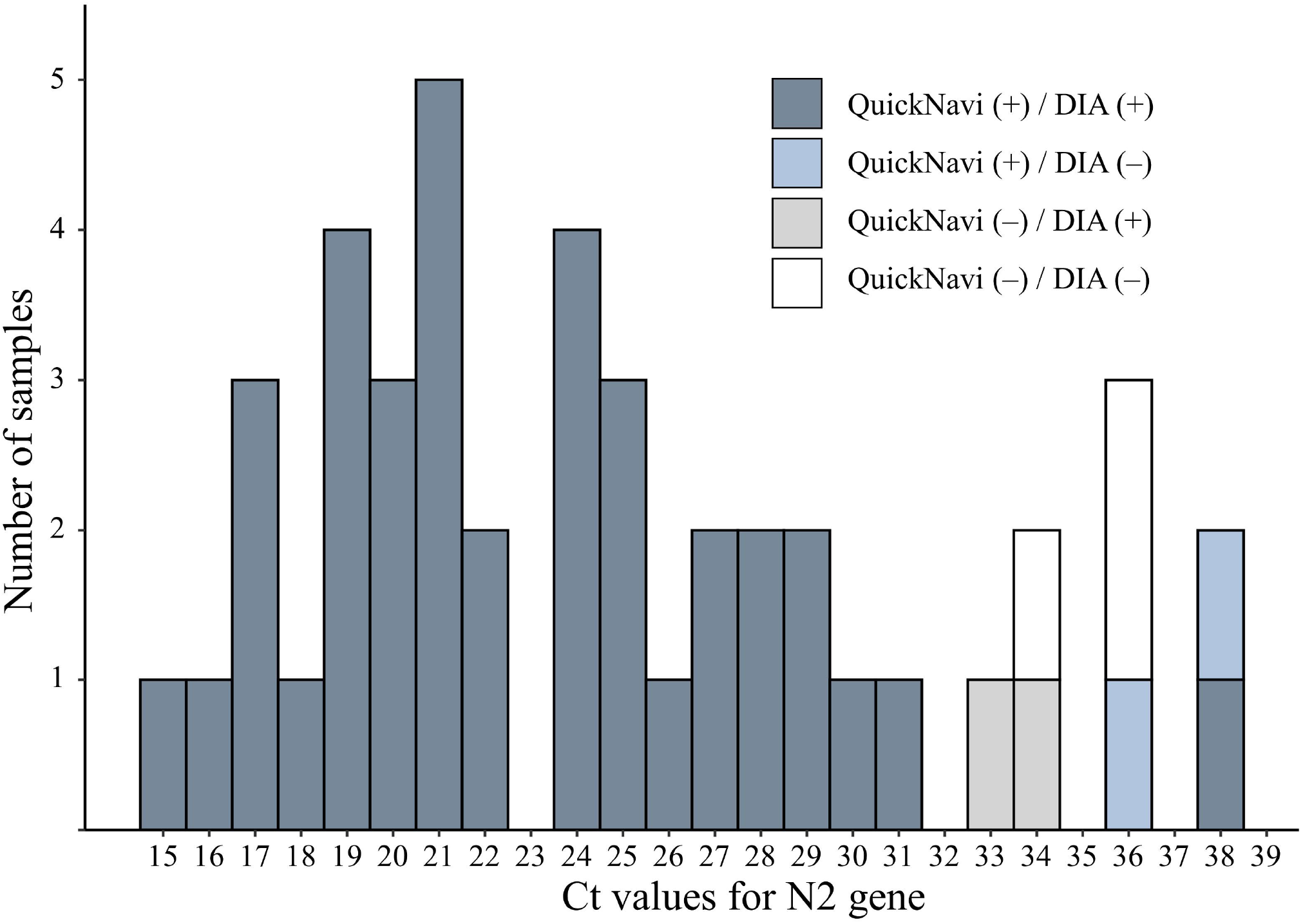
Comparison of results between QuickNavi™-COVID19 Ag and DIA. Abbreviations; QuickNavi, QuickNavi™-COVID19 Ag; DIA, digital immunoassay

## Discussion

The median time to get positive results was 15 seconds for QuickNavi™-COVID19 Ag, and 85.7% and 96.4% of patient samples resulted in positive reactions within 1 and 5 minutes, respectively. The required time to obtain positive results was significantly lengthened as the Ct values of samples increased. Direct comparison using clinical samples revealed comparable sensitivities between QuickNavi™-COVID19 Ag and the DIA.

The Centers for Disease Control and Prevention recommend that in order to avoid false negatives, the results should be read at the time designated by the manufacturer [11]. According to the manufacturer’s instruction for use, however, QuickNavi™-COVID19 Ag can be interpreted as positive once the positive line is visualized even before the specified read time. In our study, about 80% of positive cases can be tested within 1 minute and over 90% within 5 minutes, which is faster than designated read time. The manufacturer’s specified time, therefore, may be longer than necessary (Figure 3). This finding might apply to other antigen tests, although variations in performance possibly exist due to methodological differences among products [12].

The time period to obtain positive results extended proportionally with the Ct value; hence, the time required to be positive may indicate the viral load in a given sample. In our study, the time needed more than 10 minutes was limited to the cases with a Ct value >30. Given that these samples are generally culture negative [13], positive lines that appears more than a few minutes may imply a low viral load with a smaller risk of transmissivity to others. Viral load is associated with disease severity [14] in addition to transmissivity. A shorter reaction time in antigen tests might predict disease severity, considering that reaction time was employed in a clinical score to predict the severity of pneumococcal pneumonia [15]. Further studies should be conducted to evaluate the clinical usefulness of measuring the time to get positive results in various antigen tests.

We expected DIA to have a higher sensitivity than QuickNavi™-COVID19 Ag because of its lower limit of detection and its implementation of a digital immunoassay system [8]. However, the two tests had almost identical sensitivity when used with clinical samples. The reactivity of antigen tests can differ between clinical and UTM^™^ samples. QuickNavi™-COVID19 Ag detected the virus in almost all clinical samples with Ct values <30 when the supplied extraction reagent was used [7,16], but the sensitivity plummeted when samples were diluted in UTM™ [17]. Measuring the limit of detection seems insufficient to verify the clinical performance of antigen tests. In this study, four samples produced discordant results between the two tests, and all of them had Ct values >30. These discrepancies can be explained by the stochastic effects involved in detecting the virus in samples with low viral loads.

This study had several limitations. First, it did not include patients who were confirmed to be negative for SARS-CoV-2 by RT-PCR. The relationship between QuickNavi™-COVID19 Ag read times and the possibility of false positives was not evaluated, although we previously confirmed false positives are merely seen with this test [7]. Further, we could not compare the specificities of QuickNavi™-COVID19 Ag and the DIA evaluated. Second, we did not assess whether differences in strains or sample characteristics (e.g., viscosity, blood inclusion) influenced the time to get positive results.

In conclusion, QuickNavi™-COVID19 Ag yielded a positive result immediately after sample placement in most cases, and the time needed for final interpretation was longer in samples with larger Ct values. The sensitivity was comparable to that of the DIA.

## Data Availability

The data that support the findings of this study are available from the corresponding author upon reasonable request.

## ACKNOWLEDGEMENTS

We thank all staff at the Department of Clinical Laboratory of Tsukuba Medical Center Hospital who contributed to this study, and all medical institutions that provided their patients’ clinical information.

## Figure Legends

**Supplementary Figure**. Positive correlation between the time to get positive results obtained using QuickNavi™-COVID19 Ag and DK20-CoV-8M.

