## Supplementary figures and images for "Evaluation and clinical implications of the time to a positive results of antigen testing for SARS-CoV-2"

### Supplementary Figure 1

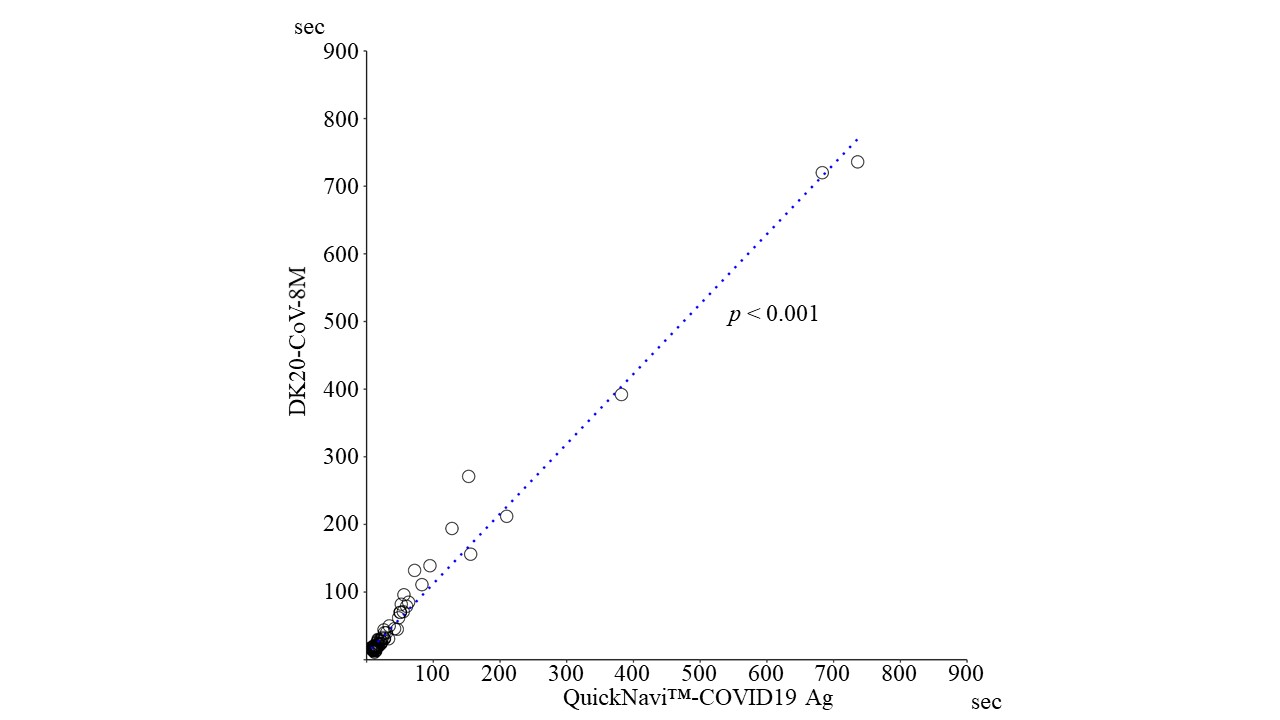
